# A Hybrid Machine Learning Model for Estimation of Obesity Levels

**DOI:** 10.1101/2022.08.17.22278905

**Authors:** Akash Choudhuri

**Affiliations:** University of Iowa, Iowa City, Iowa- 52246, United States of America

**Keywords:** Obesity, Extremely Randomized Trees, Multilayer Perceptron, XGBoost

## Abstract

Obesity has always been a problem which has plagued humans for many generations, which, since the 1975, almost doubled to turn into a global epidemic. The current human dependence on technology has contributed to the problem even more, with the effects visibly pronounced in late teenagers and early adults. Researchers till date, have tried numerous ways to determine the factors that cause obesity in early adults.

On that frontier, our hybrid machine-learning model uses the help of some supervised and unsupervised data mining methods like Extremely Randomized Trees, Multilayer Perceptron and XGBoost using Python to detect and predict obesity levels and help healthcare professionals to combat this phenomenon. Our dataset is a publicly available dataset in the UCI Machine Learning Repository, containing the data for the estimation of obesity levels in individuals from the countries of Mexico, Peru, and Colombia, based on their eating habits and physical condition. The proposed model heavily utilizes feature engineering methods and introduces the concept of a hybrid model.

This work has shown improved results over prior works and extensive studies have been undertaken to preserve the robustness of this model.

## 1. Introduction

The concept of obesity is a relevant topic in the modern society. With the advent of modern technology, mankind is trying its best to eradicate the consequences of unhealthy lifestyle, which has led to be the major contributor towards obesity. In fact, the problem of obesity prevalently grew exponentially in sync with the advancement of modern technological tools. With its origins tracing back up to 30000 years [1], even before the origin of some of the Ancient Civilizations, obesity has extensive mentions in early literature with Hippocrates mentioning how obesity led to infertility and early death.

Over time, obesity had various implications in the human society. While in some civilizations, obesity was a valuable quality to have, indicating wealth and reach, most other civilizations tried to understand the dangers obesity brought with it in the long term. However, the degree of acceptance of obesity in the human society has been comparatively slow over other major diseases. The Ancient Greeks and Egyptians seemed to despise the phenomenon. The Egyptians seemed to consider diet primarily as a source of preservation of health with texts suggesting that they vomited to prevent excess weight gain [2].

Over time, medical research seemed to align towards a more modern method to tackle obesity. This was also due to increase in the number of obese people in the society. With philosophers and writers in the 18th Century realizing preservation of health is a major reason to avoid obesity, George Cheyne MD was one of the pioneers who recognized obesity as a disease. He also stated that obesity was linked with depression [3]. One of the chief comorbidities of obesity is diabetes, which was first illustrated by Ebers Papyrus as a condition which caused ‘excessive urination’ [4]. Over time, obesity was also later linked to a multitude of conditions including coronary heart diseases and abdominal fat.

At the current time, studies in 2013 have shown that in USA, the presence of obesity is significantly vast [5].

Although researchers have been able to detect the contributors to obesity over time, however, they are yet to discover and develop foolproof methods to try to detect obesity at the very onset to combat the phenomenon. With the current developments in the domain of Machine Learning, one can construct contemporary investigative models to design accurate and precise programming routines at low computational cost, which would optimally involve a quality compromise among the typical method features. Obesity has serious consequences on our cardiovascular systems. However, the condition seems to develop in the intersection of late childhood and early adulthood. On the segment of obesity in children Cecil et al. showed that metabolic disorders which lead to obesity are equally pronounced in children as that in case of adults [6]. However, very little research has been performed on the samples of data concerning early adults. This could probably be due to the lack of resources for data collection in this segment. The current need for research on this topic is extremely important as the current Covid-19 Pandemic has affected the group of late teenagers and early adults the most, with Undergraduate and Graduate University students being highly susceptible to obesity due to online education and lack of avenues of physical activity in Covid-19 restrictions in public places [7]. So, relevant research consisting of this age group of individuals is of utmost importance.

On that note, the current research extends previous works of research in improving the accuracy of predictions of obesity estimation of young adults. Although various modern applications are available for BMI calculation, the novelty of this work lies in the fact that it not only considers weight to be a contributor of obesity. It also considers the effect of an individual’s lifestyle habits (ie, smoking habits, consumption of vegetables, quantity of water drank, modes of transportation frequented, etc.). This allows the provision for the proposed ML Model to learn representations which lead to obesity which may not be obvious to the naked eye. On that note, this study chiefly implements techniques in data mining, particularly based on multi-class classification problem, which is a subset of supervised learning to predict whether a person suffers from obesity, given a few habits and health details. The work has been divided into the following sections: Section 2 elaborates the prior research in this domain which motivated this research while Section 3 elaborates the hybrid machine learning model and the results, with special emphasis on Covid-19 and Section 4 illustrates the conclusions derived from the study.

## 2. Related Works

Previously, most of the related works in this domain have focused on trying to determine the factors that caused obesity. However, of late, with the advent and popularity of Machine Learning Algorithms and software, the process of trying to estimate obesity level has also become a major topic of research.

Amongst the prior works, the authors of [8] estimated the percentage of overweight children aged between 2 to 17 years and thus proposed an appropriate predictive model methodology. The authors of [9] used Fuzzy Signature to interpret the trends in child obesity and proposed a method to detect and handle child obesity at an early stage. In [10] general pediatric obesity development pattern and the onset time of early childhood obesity was identified along with which and XGBoost model was developed to predict if individuals have early obesity.

The authors of [11] examined childhood obesity by collecting data from a variety of sources and identified a few risk factors contributing to childhood obesity and proposed a model to predict childhood obesity using NBtree.

In [12], the authors predict childhood obesity by using only data of children collected before their second birthday. They use Weka to conclude that the Decision Tree model implemented by the ID3 algorithm performs the best in predicting childhood obesity. Meanwhile, the authors of addressed the unsupervised multi-instance learning problem by using three different kernel functions in 2 different situations, namely over the year prediction and over many counties’ prediction of health indices.

In [14], the authors used data mining to classify obesity levels among 6-year-old school children by obtaining data from 2 sources and preprocessed the resulting data and used various classification algorithms, namely Bayesian Network, Decision Tree, Neural Networks and Support Vector Machine (SVM) and a comparison was made between various feature selection techniques. The authors of [15] offered data for assessing obesity levels in persons from a few Latin American countries based on eating habits and physical condition. The synthetic data which was generated mimicking the original data of less volume have been used for many future studies on this frontier. In fact, the dataset used in the current work is the same dataset.

The authors of [5] claimed that the state level estimates from the CDC underestimate the levels of obesity in various regions in USA and further proposed a method to remove the so-called biasness in estimation. In [16] the authors state that outside of laboratory/clinical settings, current methods of self-monitoring kilocalorie intake suffer from systematic underreporting bias. During Training multiple linear regression explained if physical attributes of a person could predict an individual’s mean kilocalories per bite.

In [17] the authors proposed and compared a SVM Model with a Decision Tree model using the obesity dataset generated from the University students of Latin American countries and achieved high precision and recall values using the decision tree. Further, they proposed a Decision Tree + Simple K-Means model, where they achieved 98.5% precision and 98.5% recall. We have used this model as a baseline model to evaluate our results.

Although the previous works provide a wide array of techniques and machine learning models to predict obesity levels, most of the prior works are heavily dependent on the calculation of Body Mass Index (BMI), which does not consider other factors like hereditary history of obesity, lifestyle habits like frequent water intake, consumption (or lack of) fast foods, etc. However, with BMI. On that note, this work asserts on the development of such an intelligent tool that models obesity in human beings.

The novelty of this work can be elaborated as follows:

- This work employs extensive use of feature engineering and data preprocessing techniques which previous similar works had ignored.
- Previous works like [15] and [17] used Weka, which is just a tool. By appropriately deploying our proposed model from scratch using appropriate programming language, this work achieves greater flexibility in tuning the model and inference behind the results.
- The proposed model is extremely robust, which has been elaborated by the Covid-19 scenario testing.

## 3. Preliminaries

In this section we will discuss the concepts related to this paper.

### 3.1 Data Scaling

To place all features on the same level in many machine learning approaches, we must scale so that a large number does not harm the model simply because it is large. Feature scaling in machine learning is one of the most critical processes in the pre-processing of data prior to creating a machine learning model. Objective functions in some machine learning algorithms do not work correctly without normalization since the range of values in raw data varies substantially. The following are some ways that have been used:

- **Standardization:** Most Machine Learning Models need dataset standardization. If the individual characteristics are not like ordinary regularly distributed data, they may behave badly (generally represented between a z-score of 0 to 1). However, we disregard the distribution’s structure and transform each observation by subtracting the mean value of the feature column and dividing by the standard deviation.
- **Maximum Absolute Value Scaling:** The characteristics are scaled by their maximal absolute value in a similar manner.
- **Outlier Data:** If the data has many outliers, scaling using the mean and variance of the data gives poor performance. In such cases, RobustScaler removes the median and scales the data by interquartile range.

### 3.2 Feature Selection Methods

The number of input features/variables of a dataset which is fed into a model is generally referred to as the dimensionality of the dataset. More input features give difficulty in modelling a predictive modelling problem as it works at a higher dimensional space (curse of dimensionality). So, one needs to take a call on the exact number of input dimensions that must be fit into a model. This work has extensively employed the usage of feature selection to improve its performance.

Amongst the various dimensionality reduction and feature selection methods, we will highlight two methods that have been used in this work. They are:

- **Recursive Feature Elimination (RFE):** Recursive Feature Elimination (RFE) is a wrapper-type feature selection algorithm generally containing a core model beneath the wrapper.
- As our core machine learning algorithm in our RFE, we employed Extremely Randomized Trees discovered by Geurts et al. [18]. This choice was because ensemble approaches outperform Decision Trees in most cases and Extremely Randomized Trees outperformed Random Forest during hyperparameter tuning. This can be asserted to the commonly known fact that Extremely Randomized Trees adds randomization but still has optimization over Random Forest.
- **SelectFromModel**: This is a feature selection meta-transformer that is used with any estimator that gives priority to each feature via a specified property (weights).
- If the matching relevance of the feature values is less than the supplied threshold parameter, the characteristics are considered insignificant and eliminated. We employed the same Extremely Randomized Trees as our tree-based estimator as in the meta-transformer to compute impurity-based feature significance, which can then be used to eliminate unnecessary features.

### 3.3 Multilayer Perceptron

The multilayer perceptron (MLP) model (also known as artificial neural networks) is simply an updated form of the Rosenblatt Perceptron Model [19] in terms of design. When compared to traditional machine learning models, the multilayer perceptron model is extremely beneficial for tackling complicated issues, especially when large amounts of data are available. The design of the multilayer perceptron model is heavily influenced by the anatomy of the human brain. This allows the model to address exceedingly complex and non-linear issues that the Rosenblatt Model could not.

A multilayer perceptron model is made up of core structural components known as neurons (also called nodes). During the network’s learning process, the weighted connections between neurons can be changed, and the activation function determines the output of each neuron given a set of inputs. The neurons organize themselves into layers, and a typical multilayer perceptron design comprises of an input layer that receives the provided feature information, a few hidden layers, and an output layer that provides the model’s response. A distinguishing aspect of the MLP model is that each neuron in one layer is coupled to another neuron in the next layer.

The basic features of a multilayer perceptron model according to [20] are:

- Each neuron in the model contains a nonlinear and differentiable activation function.
- The model contains one or more hidden layer(s).
- The layers of the model are well connected, and the degree of connectivity is represented by the weights of the network.

Our work uses a fully connected MLP and the network’s signal flow proceeds in a forward direction. In our proposed model, the MLP acts as an embedding function that maps the input features into a representation in the embedding space.

### 3.4 XGBoost

As mentioned in the introductory paper by Chen and Guestrin [21], XGBoost is a scalable Tree Boosting System.

The XGBoost package implements the gradient boosting decision tree method. Boosting is an ensemble strategy in which new models are added to rectify faults generated by current models. Models are introduced in a logical order until no more developments are possible.

The XGBoost algorithm’s main benefit is that it can penalize complicated models by regularizing both L1 and L2. Regularization aids in the avoidance of overfitting. Furthermore, XGBoost may make advantage of several CPU cores for quicker computation. This function is also useful for locating column divisions and sub-samples.

As a result, XGBoost is a very beneficial performance enhancing approach. In our model (as demonstrated later), XGBoost significantly improves the model’s accuracy, allowing it to outperform the performance metrics of earlier works of study. In fact, we have demonstrated that attribute by performing an ablation study on the components of the proposed model.

## 4. Methodology

In the following section we would elaborate on the problem statement and the proposed model architecture and the functionality of each component.

### 4.1 Problem Formulation

This dataset used in this work has been obtained from the popular UCI Machine Learning repository [22]. As our job is to create a model that will determine the obesity condition of a particular individual based on the individual’s habit, we are dealing with a multi-class classification model.

The dataset was based on a study performed which was used as a primary source of the data collected from a set of students of institutions of Colombia, Mexico, and Peru. The students were aged between 18 to 25 years. The dataset description is given in the appendix.

The Data was then labeled using the formula for BMI, given by:

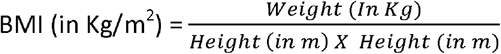

Upon identification of the balanced class problem, the tool Weka and SMOTE filter proposed by [24] produced synthetic information up to 77 percent of the data. Finally, 2111 records for each category were generated. Our classification problem was a 6-class classification problem.

### 4.2 Proposed Model Architecture

As the theoretical backgrounds about the Machine Learning Techniques, Feature Extraction and Feature Scaling has been elaborated in the previous section, we are free to refer to them to explain the model architecture.

As we observed prior works using this dataset, [15] had used decision trees using the Weka software and achieved high precision and recall values using the J48 algorithm. In our work, the shallow multilayer perceptron tries to create an embedding, which is then passed into the XGBoost model. This gives us better performance than the prior works. Figure 1 is a schematic representation of the proposed model architecture.

**Figure 1:**
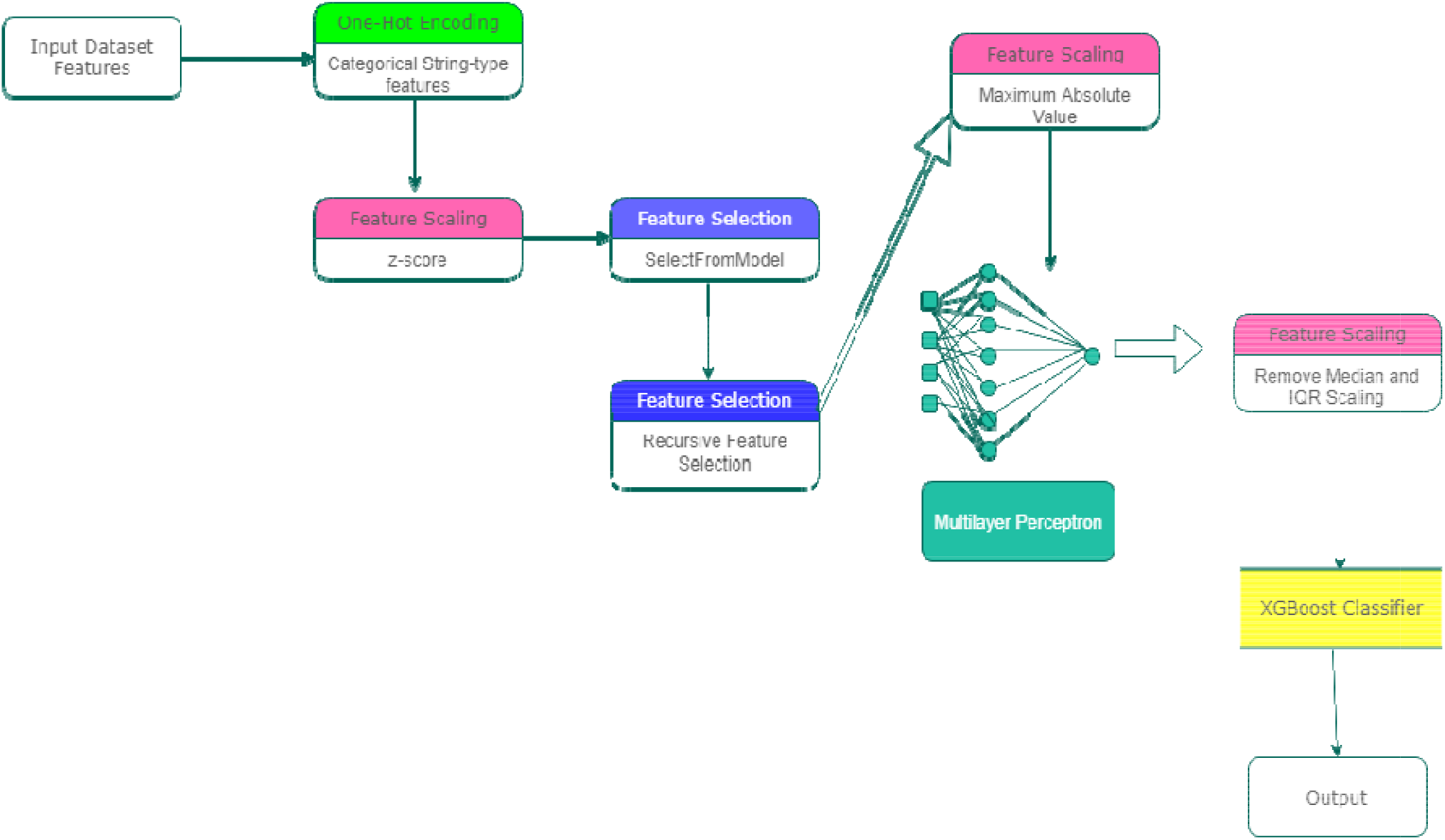
Proposed Model Architecture.

If X represents the matrix of the input training dataset and X where n and d represent the number of rows and columns respectively, then:

(1)

Here, H where m<<d and the One-Hot Encoding (a popular method to obtain a vector representation of categorical features), feature scaling and feature extraction methods are collectively referred to as Feature Engineering and is denoted by.

The dimension reduced matrix H is then passed through a shallow feed-forward Multilayer Perceptron (MLP) to generate an embedding given by:

(2)

Here 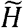 is the generated embedding and 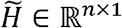. W is a learnable parameter of the Multilayer Perceptron. Then This embedding is passed through the XGBoost Classifier to generate the predictions given by:

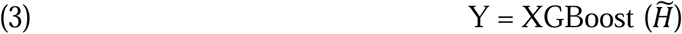

A detailed description of model parameter values is given in the Appendix.

## 5. Results and Discussion

In this section, we will discuss about the performance of the proposed model based on metrics and compare our results to previous works. Further, we would also ascertain the importance of the XGBoost model in this architecture and examine the robustness of the model predictions in the context of external stress (Covid-19).

### 5.1 Experimental Setup and results

To train our model, we used the training data consisting of 1583 samples and trained out hybrid model. Then we tested our model using the test data. Our performance metrics, like [25], which also worked with the multi-class classification problem, were as follows:

a. **Confusion Matrix:** This is a matrix which gives a comparative analysis between the ground truth values and the values predicted by our model. As our problem is a six-class classification problem, the confusion matrix in this case is a 6 × 6 matrix. In the confusion matrix, we get the counts of **True Positives (TP)** or the number of times the model correctly predicted the positive ground truth, **True Negatives (TN)** or the number of times the model correctly predicted the negative ground truth, **False Positives (FP)** also known as Type I error and **False Negatives (FN)** also known as Type II error.
b. **Average Accuracy:** It is formulated as:

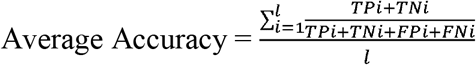
c. **Precision:** In a multi-class classification problem, precision is calculated in both micro (Precision_μ_) and macro (Precision_M_) level. The formulae are given by:

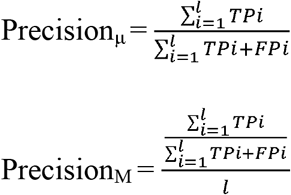
d. **Recall:** Like precision, multi-class classification has micro (Recall_μ_) and macro (Recall_M_) recall. The formulae are given by:

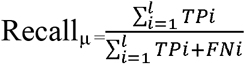

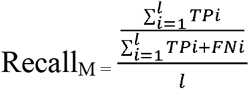

The confusion matrix of the model is demonstrated in Table 1.

**Table 1:**
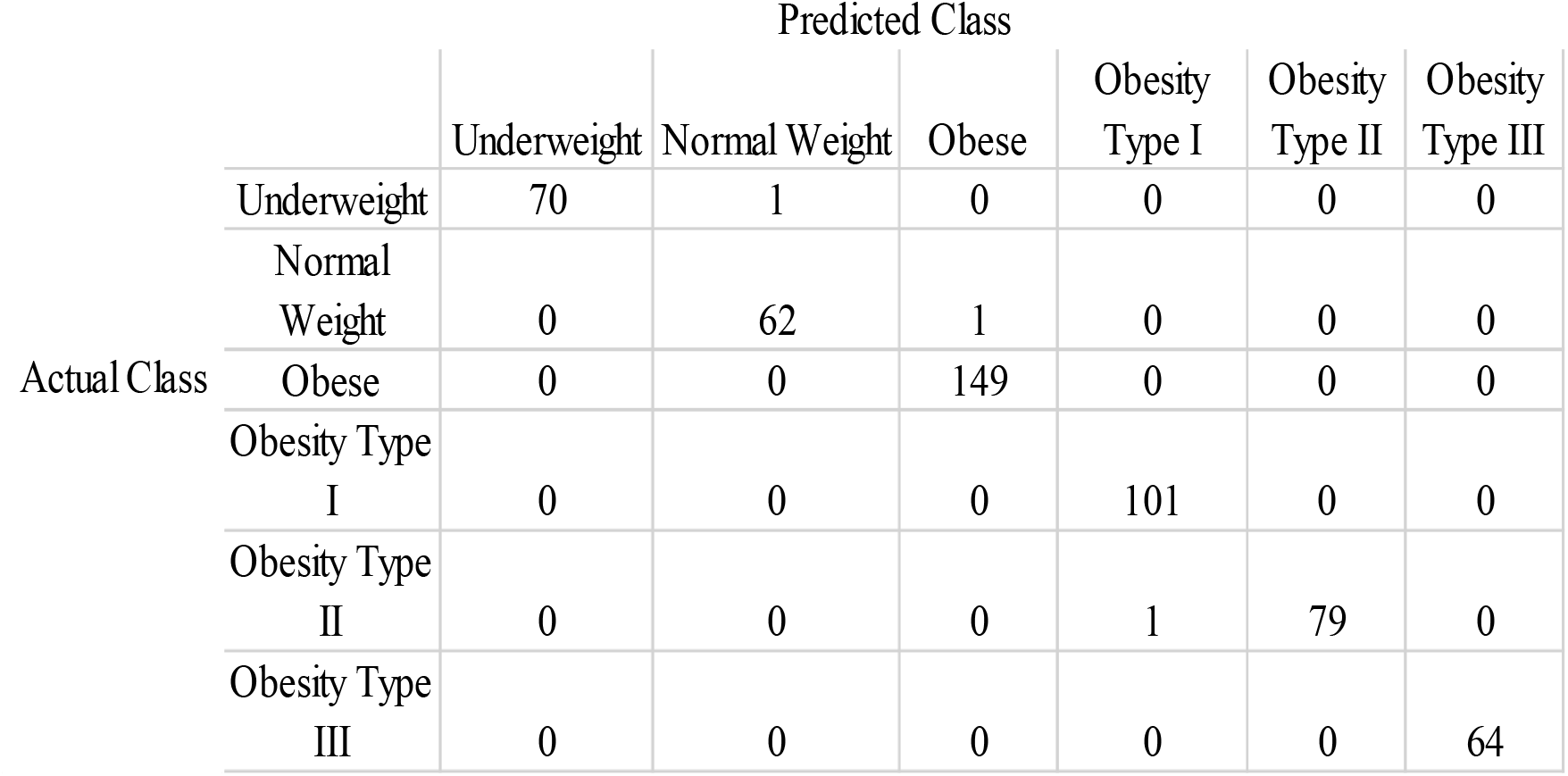
Confusion Matrix for the 6-class classification Problem.

The values of the different performance metrics are given is Table 2.

**Table 2:** Performance metric values of the proposed model.

| Performance Measure | Value |
| --- | --- |
| **Accuracy** | 0.994318 |
| **Precision<sub>μ</sub>** | 0.994349 |
| **Recall<sub>μ</sub>** | 0.994318 |
| **Precision<sub>M</sub>** | 0.994609 |
| **Recall<sub>M</sub>** | 0.992923 |

The model performance for each individual class can is elaborated in the Appendix.

### 5.2 Comparison of Results to those of Previous Works

We had previously mentioned that the Decision Tree model proposed [15] is our baseline research model. Another similar work with better results were proposed by [17]. Thus, we will try to compare our results to the results reported by the authors. There was no way to validate or verify their results and so we are assuming that the values reported were true values. Table 3 shows the comparison of the results. We notice that the proposed model outperforms each of the previous models in both performance metrics.

**Table 3:** Comparison of results of the proposed model with previous works.

| Model Name | Precision | Recall |
| --- | --- | --- |
| **Decision Tree (J48) [15]** | 97.4 % | 97.8 % |
| **Decision Tree + Simple K-Means [17]** | 98.5 % | 98.5% |
| **Hybrid Current Model** | **99.4 %** | **99.4 %** |

**Table 4:** Performance of the Hybrid model in COVID-19 simulation.

| Performance Measure | Value |
| --- | --- |
| Accuracy | 0.990530 |
| Precision<sub>μ</sub> | 0.990674 |
| Recall<sub>μ</sub> | 0.990530 |
| Precision<sub>M</sub> | 0.991474 |
| Recall<sub>M</sub> | 0.983301 |

So, this work reasonably outlines the usage of hybrid machine learning models in this domain of obesity detection in human. Also, the current model also significantly urges the researchers and practitioners in the domain to explore more into the currently newer performance enhanced models available in the industry. Also, this work also shows how important data pre-processing steps are. The whole process of frequent feature extraction and feature scaling were pivotal in improving performance.

### 5.3 Ablation Study

To address possible multi-class classification characteristics, various machine learning algorithms were used. Amongst all the combinations, we discovered that the Multilayer Perceptron + XGBoost gave the highest performance. However, it must be remembered that this performance was only with respect to the dataset provided which captures the obesity dynamics of a particular region for a particular age group. Thus, due to the lack of generalization in the given dataset and lack of publicly available data sources on this domain, it was not possible to provide a model that will predict correctly for all general cases.

Using different varieties of machine learning methods proved to be very effective in improving the performance of the model. On running a diagnostic, we observed that the models, in a stand-alone condition, performed well. But their performance rose rapidly, when included in the pipeline, one after the other. In the experiment, we tried to test for the independent prediction power of the XGBoost model without the MLP Model. However, we have used all the dimensionality reduction methods and feature standardization methods as previously mentioned in the pipeline. Thus, the embedding is an important part of the model. Figure 2 shows the result.

**Figure 2:**
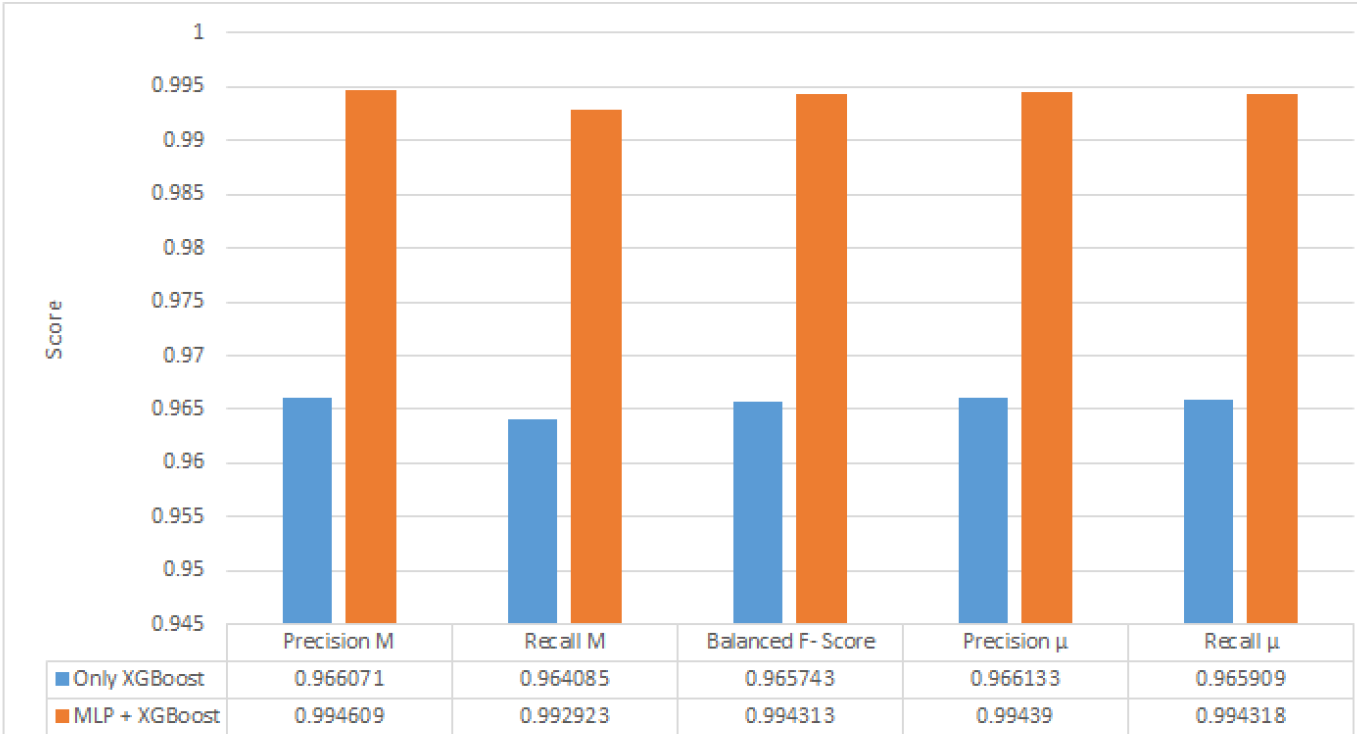
Result of the ablation study.

### 5.4 Study Involving COVID-19

We decided to stress-test our hybrid model architecture on the onset of Covid-19. As we already know, the given data was taken much before Covid-19. So, we decided to use the findings of [7] and assumed the period of Covid-19 to be on peak and lockdowns in various countries to be actively during the time Covid-19 peaked in Latin America. On 20^th^ March 2020, the Columbian President had declared an initial 19-day lockdown from 24^th^ March 2020, which was extended up to 27^th^ April 2020, being again extended to 31^st^ May 2020, finally going up to 1^st^ September 2020. Lockdown was declared in Peru from 16^th^ March 2020 to 8^th^ September 2020. Venezuela also went into lockdown from 17^th^ March 2020 up to July 2020.

Thus, we are considering the period of lockdown due to Covid-19 to be held from 20^th^ March 2020 up till 1^st^ September 2020 and using the value of average weight gain mentioned in [7] by initializing a random multiplier between 0-0.27Kg of every 10 days for the given period. Further details of this study are mentioned in the appendix. The performance of the model is, as follows.

## 6. Conclusions

In this work of research, we introduced a hybrid Machine Learning Model for Human Obesity Detection using various underlying statistical and machine learning concepts. In our model pipeline, we utilized a plethora of dimensionality reduction and feature scaling methods, which were very important in maximizing the performance of predictions of our said model. Also, we employed adequate amount of time and effort in performing hyper parameter tuning for all the internal constituents of this overall model structure.

We employed the usage of two widely popular prediction supervised learning methods, namely the Multilayer Perceptron Model (also known as Artificial Neural Network) and the tree-based ensemble technique XGBoost. Also, we employed judicious usage of Extremely Randomized Trees Classifier as the tree-based dimensionality reduction method, both in Recursive Feature Elimination and SelectFromModel.

Our constructed model gave a prediction accuracy of 99.43%, a precision value of 0.994 and a recall value of 0.994, which were sizeable improvements from the performance metrics generated from previous works of research working with the same dataset. We also tried to estimate the values of the different performance metrics by creating a simulated dataset keeping the onset of Covid-19 in mind and used the same model parameters and pipelines to validate the power of predictions of the model. Unsurprisingly, we did not find any major discrepancy in this experiment, with the model architecture reporting accuracy of 99.05%, precision score of 0.990 and a recall score of 0.990.

An extension towards the future scope of this work could be extended in attempting to improve the accuracy of the classification with wider spectrum of data covering candidates from more countries and inclusion of more ML methods into the model pipeline. This approach would result in a more fine-grained obesity classification as with the passage of time, as we are making bigger breakthroughs in Machine Learning research and algorithms.

Another scope of improving the current work would be further optimization of the parameters of the Machine Learning techniques used in the pipeline as some of the model parts could not be optimized fully due to limitation of resources. In fact, in just the Multilayer Perceptron (Artificial Neural Network), further hyperparameter tuning could provide better estimates of globally optimized values, which will improve the state of predictions of the model. In addition to that, efforts can be made to present more publicly available datasets on obesity, which would be homogenous in terms of the features and descriptions. This would harbinger a newer direction of research in trying to build a model that will be able to identify the global trend as a whole and make accurate predictions.

To conclude, the given work discusses a few popular Machine Learning Techniques and combines them to create a hybrid Machine Learning Model for the classification of levels of obesity which significantly outperforms previous works of research.

## Data Availability

All data produced are available online at https://archive.ics.uci.edu/ml/datasets/Estimation+of+obesity+levels+based+on+eating+habits+and+physical+condition+

https://github.com/Soothysay/HybridObe

## Acknowledgements

The author would like to place on record their sincere thank Prof. Sudarsan Padhy for his valuable insight in improving the quality of results and guiding the author regarding the theoretical explanations mentioned in this work. The author offers his gratitude to the management and staff of Institute of Mathematics and Applications Bhubaneswar, for their constant support.

## Declaration by the Author

The author would like to hereby declare that the above work is an original work, and the paper has not been submitted or accepted for publication anywhere else. The author’s contribution is the construction of the proposed Hybrid ML Model using Python 3.7 from scratch.

