## Supplement for "A Hybrid Machine Learning Model for Estimation of Obesity Levels"

**Appendix I: Original Dataset Description**

The Dataset used in this work is publicly available at <https://archive.ics.uci.edu/ml/datasets/Estimation+of+obesity+levels+based+on+eating+habits+and+physical+condition+>.

The code is publicly available at <https://github.com/Soothysay/HybridObe> .

The Dataset Description is:

| Attributes | Values |
| --- | --- |
| Gender | - Male - Female |
| Age | Numeric value in years. |
| Height | Numeric Value in metres. |
| Weight | Numeric Value in Kilograms. |
| Family Member Overweight | - Yes - No |
| Regular High Calorie Food Intake | - Yes - No |
| Consumption of Vegetables in Meals | - Never - Sometimes - Always |
| Number of Daily Main Meals Intake | - Between 1-2 - 3 - More than 3 |
| Food Intake between Meals | - No - Sometimes - Frequently - Always |
| Smoke | - Yes - No |
| Daily Water Intake | - Less than 1 litre - 1-2 litre - More than 2 litres |
| Calories Consumption Calculation | - Yes - No |
| Physical Activity in a Week | - No physical activity - 1-2 days - 2-4 days - 4-5 days |
| Alcohol Consumption | - Non drinker - Sometimes - Frequently - Always |
| Schedule Given to Technology | - 0-2 hours - 3-5 hours - More than 5 hours |
| Mode of Transportation Used | - Automobile - Motorbike - Bike - Public Transportation - Walking |

**Appendix II: Model Parameter Values**

Extensive hyperparameter tuning was used. The codes were run with Python 3.7 on a machine equipped with i5-8265U processor and 8 GB RAM. The code took 1.3s to execute. The model parameter values used were:

| Model Part | Parameter Values |
| --- | --- |
| Feature Selection (Select From Model) | Extremely Randomized Trees Classifier:   - Number of trees in forest = 100. - Criterion = Gini. - Maximum Number of Features to consider best split = 0.1 |
| Feature Selection (Recursive Feature Elimination) | Extremely Randomized Trees Classifier:   - Number of trees in forest = 100. - Criterion = Gini. - Maximum Number of Features to consider best split = 0.4 |
| Multilayer Perceptron Classifier | - Number of hidden layers = 1. - Number of neurons in hidden layer = 100. - Optimizer = Adam (Learning rate = 0.5). - L2 Regularization Parameter value = 0.1. |
| XGBoost Classifier | - Learning Rate = 0.5. - Maximum Depth of a Tree = 7. - Minimum sum of instance weight (hessian) needed in a child = 1. - Subsample ratio of the training instances = 0.8500000000000001. |

**Appendix III: Model Performance for Each Individual Class**

| Class | Accuracy | Precision | Recall | f1- Score | Support |
| --- | --- | --- | --- | --- | --- |
| Underweight | 99.81% | 100% | 98.59% | 99.29% | 71 |
| Normal Weight | 99.62% | 98.41% | 98.41% | 98.41% | 63 |
| Obese | 99.81% | 99.33% | 100% | 99.66% | 149 |
| Obesity Type I | 99.81% | 99.01% | 100% | 99.50% | 101 |
| Obesity Type II | 99.81% | 100% | 98.75% | 99.37% | 80 |
| Obesity Type III | 100% | 100% | 100% | 100% | 64 |

**Appendix IV: Additional Material for COVID-19 Study**

The details of the COVID-19 stress testing study is given as follows:

| Parameter | Value/ Usage |
| --- | --- |
| Start Date | 20/3/2020 |
| End Date | 1/9/2020 |
| Total Duration in days | 165 |
| Random Multiplier Value | Increase of [0, 0.27] Kg weight for every 10 days. |

The resultant weight histogram is shown as below and compared to the previous value in Figure A and Figure B. In fact, the general trend of weights has also changed significantly.

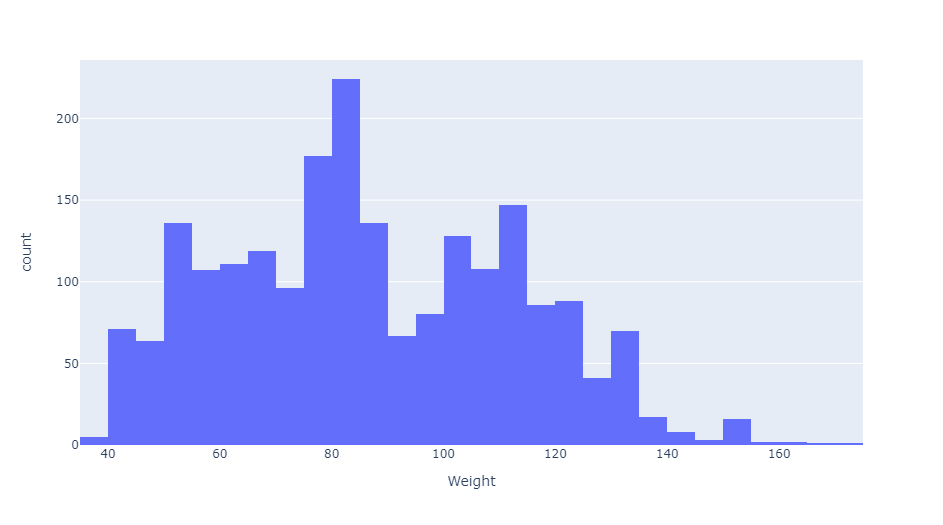

**Figure A:** Histogram of Weights before Covid-19.

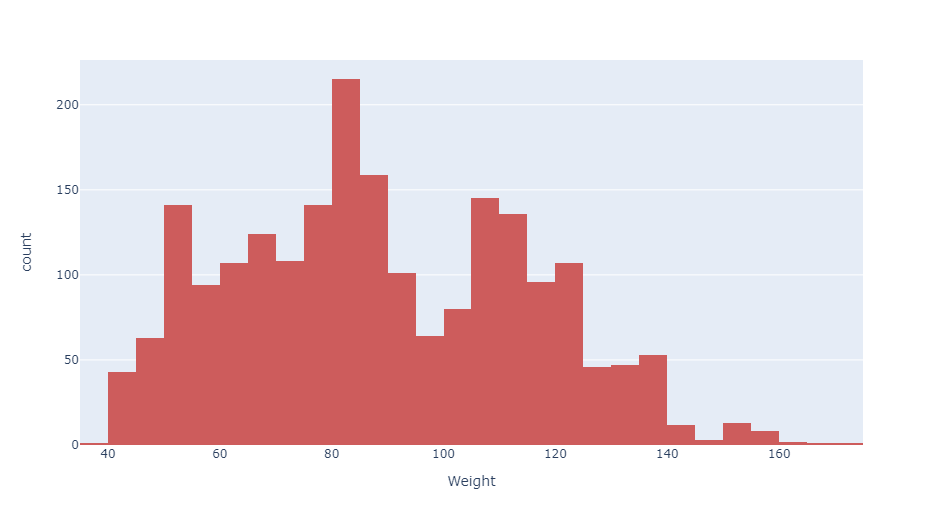

**Figure B:** Histogram of Simulated Weights after Covid-19.
